# Anatomic Predilection of IDH-Mutant Gliomas: A Multi-Institutional Spatial Analysis

**DOI:** 10.1101/2025.09.16.25333605

**Authors:** Minjun Park, Hannah Weiss, Edward S. Harake, Camila Fang, Alex Springer, Nicholas K. Goff, John E. Markert, David Reinecke, Nader Maarouf, Alex M. Miller, Todd Hollon, John G. Golfinos, Daniel A. Orringer

**Author notes:** Corresponding author: Daniel A. Orringer, MD Department of Neurological Surgery, New York University Langone Medical Center 530 First Avenue, Skirball Suite 8R, New York, NY 10016. These authors contributed equally to this work (Minjun Park, Hannah Weiss). Funding Disclosure Statement: No financial support for this study was received.

## Abstract

**Introduction:** Interactions between cancer cells and their microenvironment are central to tumor formation. Regional microenvironmental variability in the brain may offer insight into essential factors in tumorigenesis. Surprisingly, a granular assessment of regional patterns of gliomagenesis has not been undertaken in the molecular era.

**Objective:** To quantitatively establish the anatomical distribution of the major molecular subtypes of adult diffuse glioma.

**Methods:** We retrospectively analyzed a consecutive series of 204 IDH-mutant and 200 IDH-wildtype gliomas. Tumor volumes were segmented, normalized, and assigned to a brain location using a standardized neuroanatomic atlas. An independent and external validation cohort of 190 IDH-mutant and 227 IDH-wildtype gliomas was used to assess reproducibility. Microarray expressions from the Allen Human Brain Atlas were utilized to analyze transcriptomic differences between hotspots and coldspots of IDH-mutant gliomas.

**Results:** 50.5% (103/204) of IDH-mutant tumors arose with the superior and middle frontal gyri indicating a 3.1-fold regional enrichment relative to the volume of these gyri (p<0.001). Only 0.5% (1/204) of IDH-mutant tumors arose with the occipital gyri indicating a 23.4-fold regional scarcity relative to volume (p<0.001). 9.5% (19/200) of IDH-wildtype tumors arose in the superior temporal gyrus with a 2.1-fold enrichment (p=0.01) and 6% (12/200) in the hippocampus with a 6-fold enrichment (p<0.001). IDH-mutant and wildtype tumors were enriched by 4 and 4.5-fold respectively in the insula (both p<0.001). Both IDH-mutant astrocytomas and oligodendrogliomas were significantly enriched in the SFG (39.2% and 40.2% respectively; both p<0.001), but 23.5% (24/102) of astrocytomas occurred disproportionately higher in the insula compared to oligodendrogliomas (p<0.001). These enrichment patterns were reproduced in an independent validation cohort of 417 glioma patients. Transcriptomic analysis comparing the lobar hotspot (frontal lobe) to the coldspot (occipital lobe) revealed frontal enrichment of cholesterol (NES=1.78) and fatty acid (NES=1.94) metabolism pathways, paralleling the observed regional enrichment of IDH-mutant gliomas.

**Conclusions:** This study reveals striking regional differences in glioma incidence, identifying key hotspots in the brain for gliomagenesis that vary based on tumor molecular subtype. Metabolic differences across cortical regions raise the possibility that regional metabolic differences may contribute to the observed vulnerability of specific regions to gliomagenesis. These findings provide a framework for investigating additional microenvironmental factors that drive human glioma formation.

## Introduction

The development and progression of gliomas are increasingly recognized as being governed not only by cell-intrinsic mutations but also by the complex interplay between neoplastic cells and their surrounding microenvironment. Accumulating evidence suggests that regional variation in glial lineages, vascular architecture, metabolic gradients, and immune architecture shape the molecular context in which gliomas emerge.^1–10^ These regionally distinct microenvironments may harbor unique transcriptional and epigenetic signatures, which may influence both tumor initiation and phenotype. However, despite advances in molecular neuropathology, the extent to which distinct cerebral territories exhibit intrinsic vulnerability or resistance to IDH-mutant gliomagenesis remains poorly defined.

Recent advances in molecular neuropathology – particularly the identification of isocitrate dehydrogenase (IDH) mutations – have redefined the biological classification of gliomas and revealed profound differences in their clinical behavior and therapeutic responsiveness.^11–14^ IDH-mutant gliomas are associated with prolonged survival and unique metabolic and epigenetic profiles, which has spurred the development of targeted therapies.^15–19^ However, the spatial predilections of IDH-mutant vs IDH-wildtype gliomas remain poorly resolved. Existing studies have been constrained by limited cohort sizes, imprecise lobar categorizations, and pre-molecular diagnostic classifications schemes.^20–23^

Defining whether certain brain regions exhibit disproportionate vulnerability to glioma formation could yield critical insights into microenvironmental factors that foster gliomagenesis. A spatially resolved molecularly stratified map of glioma origin may refine early detection algorithms, support region-informed surgical and diagnostic strategies, and illuminate context-specific therapeutic targets. In this study, we employ a high resolution cortical and subcortical atlas to systematically map the anatomical distribution of adult gliomas stratified by IDH status. We further integrate transcriptomic data from the human brain to explore whether gene expression differences contribute to regional glioma susceptibility. Together, these efforts aim to establish the first atlas based framework for uncovering spatially dependent molecular mechanisms underlying glioma pathogenesis.

## Methods

### Study Design and Patient Cohorts

We performed a retrospective, multi-institutional spatial analysis of adult diffuse gliomas across three data sources: (1) New York University (NYU) Langone Health, (2) University of Michigan Health, and (3) The Cancer Genome Atlas (TCGA).^24^ All patients were ≥ 18 years of age and had histologically confirmed WHO grade 2-4 gliomas with available genetic profiling and high-resolution pre-treatment magnetic resonance imaging (MRI). At NYU, 204 consecutive patients with IDH-mutant gliomas treated between 2015 and 2024 were included. For the control cohort, 200 consecutive adult patients with IDH-wildtype gliomas and MRI scans available at time of diagnosis were analyzed. This study was approved by the Institutional Review Board of NYU (i22-00302).

For the external validation cohort, we identified 139 IDH-mutant and 125 IDH-wildtype gliomas from the University of Michigan (2013-2023), and 51 IDH-mutant and 102 IDH-wildtype gliomas from TCGA, for a total of 821 tumors across all datasets. Clinical data-including demographics, presenting symptoms, molecular alterations, treatment details, and outcomes-were extracted for the NYU cohort.

### Imaging Acquisition and Spatial Analysis

Pre-operative or earliest available MRI images were reviewed for all patients. Tumor volumes were manually segmented by trained reviewers and validated by a second neurosurgeon. Segmentations were co-registered to standard Montreal Neurological Institute (MNI152) space. Tumor outlines were mapped to the Automated Anatomical Labeling atlas version 3 (AAL3)^25^ to assign each tumor to a dominant anatomical label. For cases overlapping multiple regions, the atlas label containing the largest tumor volume was assigned as the primary tumor site (Figure 1).

**Figure 1.** Study workflow. (A) Spatial analysis: Imaging data from TCGA, NYU, and University of Michigan cohorts were pooled and registered to MNI space. Tumor masks used existing segmentations for TCGA and manual segmentations for NYU/Michigan, which were validated and refined by the senior investigator. All segmentations were mapped to the AAL3 atlas for volumetric spatial analysis. Clinical variables were abstracted for the NYU cohort. (B) Gene expression analysis: Six microarray datasets from the Allen Human Brain Atlas were normalized and filtered; differential expression, weighted gene co-expression network analysis (WGCNA), and gene set enrichment analysis (GSEA) were performed to compare regions of interests derived from the spatial analysis.

Observed tumor frequencies by region were compared to expected frequencies derived from AAL3 regional voxel volume to identify statistically enriched or underrepresented locations. Statistical comparisons included a binomial test with a two-sided alternative and a Benjamini-Hochberg correction for p-value adjustment. Additional methodological details, including details on image preprocessing, segmentation protocol, location mapping, and data preparation for spatial analysis are provided in *Supplemental Digital Content*.

### Transcriptomic Analysis of Regional Vulnerability

We analyzed normalized microarray data from six neurotypical adult brains in the Allen Human Brain Atlas to assess regional gene expression differences associated with tumor distribution (Figure 1).^26^ Microarray samples were parcellated according to the AAL3 atlas and probes with background signals were filtered.

Differential gene expression analysis compared transcriptomic profiles between tumor-enriched and tumor-sparse regions. Genes with adjusted p < 0.05 and log fold change > 1 were considered differentially expressed.

We performed weighted gene co-expression network analysis (WGCNA), correlating gene expression with anatomical location. Gene set enrichment analysis (GSEA) was also performed on modules of interest from WGCNA and between tumor-enriched and tumor-sparse regions. Significance was defined as p < 0.05 and false discovery rate (FDR) adjusted p-values < 0.25 or < 0.05 based on the type of GSEA algorithm ran.^27^

Comparisons were made both between frontal and occipital lobes and between superior and inferior frontal gyri. Details of data processing and gene expression analysis can be found in *Supplemental Digital Content*.

## Results

We analyzed 821 gliomas, including 394 IDH-mutant and 427 IDH-wildtype tumors from two institutions and one public database.

### Clinical and Molecular Characteristics of the NYU IDH-Mutant Cohort

Among the 204 patients with IDH-mutant gliomas at NYU, the median age at diagnosis was 46.0 years (IQR 37-57), and 52.5% (n=107) were female (Table 1). Tumor laterality was evenly distributed (Table 2). IDH-mutant tumors were nearly evenly split between astrocytoma with ATRX loss (50.5%, n=103) and oligodendroglioma with 1p/19q co-deletion (49.5%, n=101).

**Table 1.** Baseline Patient Characteristics.

| Characteristics |  | N=204 |
| --- | --- | --- |
| **Sex** |  |  |
| M |  | 97 (48.1%) |
| F |  | 107 (51.9%) |
| **Age (Mean ± SD)** |  |  |
|  |  | 46.4 ± 13.3 |
| **Race** |  |  |
| White not of Hispanic American Origin |  | 128 (62.1%) |
| Black not of Hispanic American Origin |  | 4 (1.9%) |
| Hispanic |  | 24 (11.7%) |
| Asian |  | 20 (10.2%) |
| American Indian |  | 4 (1.9%) |
| Other |  | 5 (2.4%) |
| Unknown |  | 21 (9.7%) |
| **Ethnicity** |  |  |
| Not of Spanish or Hispanic Origin |  | 175 (85.0%) |
| Hispanic or Latino or Spanish |  | 26 (12.6%) |
| Unknown |  | 5 (2.4%) |
| **Presenting Symptoms** |  |  |
| Seizures |  | 103 (50.2%) |
| Cognitive Decline |  | 9 (4.4%) |
| Weakness/Motor Deficit |  | 7 (3.4%) |
| Cranial Nerve Deficit |  | 3 (1.5%) |
| Headache |  | 43 (21.0%) |
| Imbalance |  | 3 (1.5%) |
| Incidental |  | 17 (8.3%) |
| Sensory Deficit |  | 5 (2.4%) |
| Other |  | 8 (3.9%) |
| Unknown |  | 7 (3.4%) |
| **Handness** |  |  |
| Right |  | 193 (93.7%) |
| Left |  | 4 (1.9%) |
| Ambidextrous |  | 9 (4.4%) |
| **Initial Surgical Intervention** |  |  |
| Biopsy |  | 25 (12.1%) |
| Resection |  | 181 (87.9%) |
| **Extent of Resection** |  |  |
|  | Gross Total Resection | 73 (40.3%) |
|  | Near Total Resection | 33 (18.2%) |
|  | Subtotal Resection | 62 (34.3%) |
|  | Unknown | 13 (7.2%) |
| **Time to Presentation to Resection (Days, Median)** |  |  |
|  |  | 31 |
| **Number of Re-resections** |  |  |
|  | 2 | 86 (41.7%) |
|  | 3+ | 9 (4.4%) |
| **Treatment Received** |  |  |
|  | Temozolomide | 108 (44.7%) |
|  | Bevacizumab | 15 (7.3%) |
|  | IDH Inhibitors | 39 (18.9%) |
|  | Radiotherapy | 140 (29.1%) |
|  | Gamma Knife | 4 (2.4%) |
| **KPS** |  |  |
|  | 100 | 44 (24.0%) |
|  | 90 | 87 (47.5%) |
|  | 80 | 27 (14.8%) |
|  | 70 | 10 (5.5%) |
|  | 60 | 8 (4.4%) |
|  | <50 | 5 (2.4%) |
|  | Unknown | 2 (1.1%) |
| **Overall Survival (Days, Mean)** |  |  |
|  |  | 2458 |
| **Median Time to First Progression (Days)** |  |  |
|  |  | 1452 |
| **Median Time to Second Resection from First (Days)** |  |  |
|  |  | 981 |
| **Patient Status** |  |  |
|  | Alive | 192 (93.2%) |
|  | Deceased | 12 (5.8%) |
|  | Unknown | 2 (1.0%) |
| **Mean Follow-Up Time (Years, Mean)** |  |  |
|  |  | 6.7 |

**Table 2.** Tumor Characteristics and Molecular Profiling.

|  |  |  |
| --- | --- | --- |
| **Enhancement** |  |  |
|  | Yes | 125 (61.3%) |
|  | No | 59 (28.9%) |
|  | Unknown | 20 (9.8%) |
| **Significant Edema** |  |  |
|  | Yes | 150 (73.5%) |
|  | No | 33 (16.2%) |
|  | Unknown | 21 (10.3%) |
| **Laterality** |  |  |
|  | Left | 107 (52.5%) |
|  | Right | 97 (47.5%) |
| **Final Diagnosis** |  |  |
|  | Astrocytoma, IDH mutant, ATRX deletion | 103 (50.5%) |
|  | Oligodendroglioma, IDH mutant, 1p19q co-deletion | 101 (49.5%) |
| **MGMT Status** |  |  |
|  | Methylated | 124 (60.8%) |
|  | Unmethylated | 56 (27.5%) |
|  | Indeterminate | 2 (1.0%) |
|  | Not available | 22 (10.8%) |
| **Specific IDH Mutation** |  |  |
|  | R132H, R132C, R132S, R132G | 197 (96.6%) |
|  | R172K, R172W, R172M | 5 (2.5%) |
|  | Unknown | 1 (0.5%) |
| **CDKN2A Status** |  |  |
|  | Wildtype | 171 (83.8%) |
|  | Homozygous deletion | 7 (3.4%) |
|  | Hemizygous deletion | 2 (1.0%) |
|  | Amplification | 2 (1.0%) |
|  | Point mutations or promoter methylation | 2 (1.0%) |
|  | Unknown | 20 (9.8%) |
| **ATRX Status (Astrocytoma)** |  |  |
|  | Intact | 102 (50.0%) |
|  | Missing | 85 (41.7%) |
|  | Unknown | 15 (7.4%) |
|  | Other | 2 (1.0%) |
| **1p19q co-deletion (Oligos)** |  |  |
|  | Yes | 99 (48.5%) |
|  | No | 94 (46.1%) |
|  | Unknown | 11 (5.4%) |
| **TERT Promotor Mutation** |  |  |
|  | No | 129 (63.2%) |
|  | Yes | 59 (28.9%) |
|  | Unknown | 16 (7.8%) |
| **BRAF V600E Mutation** |  |  |
|  | No | 197 (96.6%) |
|  | Yes | 3 (1.5%) |
|  | Unknown | 4 (2.0%) |

### Spatial Distribution of IDH-Mutant Gliomas

IDH-mutant gliomas were strongly concentrated in the frontal lobe. Half (50.5%) arose within the superior and middle frontal gyri, representing a 3.1-fold enrichment relative to regional brain volume (*adj-p* < 0.001). The superior frontal gyrus (SFG) alone accounted for 39.7% of cases (81 observed vs 23 expected; observed-to-expected [O:E] = 3.5; *adj-p* < 0.001) (Table 3A). The middle frontal gyrus (MFG) was also enriched, although less than SFG, with 2.2 fold more IDH-mutant tumors than expected (*adj-p* <0.001). The inferior frontal gyrus (IFG) had an observed to expected ratio of 1.0 (8 observed vs 8 expected) with IDH-mutant tumors (*adj-p* = 1). Tumor incidence in the insula was 7 fold higher than expected (28 observed vs. 4 expected, *adj-p* < 0.001). In contrast, the temporal and occipital lobes were underrepresented (O:E = 0.47 and 0.04, respectively; both *adj-p* < 0.001) (Figure 2).

**Figure 2.** Spatial distribution of gliomas (NYU cohort). Dashed lines indicate expected counts proportional to atlas volume. (A) Lobar distribution: IDH-mutant gliomas are enriched in the frontal lobe; IDH-wildtype gliomas are enriched in the temporal lobe; both groups are depleted in the occipital lobe. (B) Atlas-level distribution: IDH-mutant gliomas predominantly involve the superior and middle frontal gyri; IDH-wildtype gliomas show hippocampal involvement; both groups are elevated in the insula. * Observed counts were significantly higher than expected. ** Observed counts were significantly lower than expected.

**Table 3.** Observed-to-Expected Ratios in NYU Cohort.

| Table 3A. Observed-to-Expected Ratios of IDH-Mutant Gliomas, NYU Cohort (n=204) |  |  |  |  |
| --- | --- | --- | --- | --- |
| Location | Observed | Expected | Adjusted p-value | Observed to Expected Ratio |
| Insula | 28 | 4 | < 0.001 | 7 |
| Superior frontal gyrus | 81 | 23 | < 0.001 | 3.52 |
| Middle frontal gyrus | 22 | 10 | 0.012 | 2.2 |
| Cerebellar hemispheres | 2 | 24 | < 0.002 | 0.08 |
| Table 3B. Top 5 Observed-to-Expected Ratios of IDH-Wildtype Gliomas, NYU Cohort (n=200) |  |  |  |  |
| Location | Observed | Expected | Adjusted p-value | Observed to Expected Ratio |
| Hippocampus | 12 | 2 | < 0.001 | 6 |
| Insula | 18 | 4 | < 0.001 | 4.5 |
| Thalamus | 9 | 2 | 0.01 | 4.5 |
| Superior temporal gyrus | 19 | 9 | 0.014 | 2.1 |
| Cerebellar hemispheres | 8 | 24 | < 0.001 | 0.33 |

The validation cohort demonstrated similar patterns, with the SFG remaining enriched by 2.8 fold (*adj-p* < 0.001). No significant spatial differences were noted between the discovery and validation cohorts (see *Supplemental Digital Content*).

### Spatial Distribution of IDH-Wildtype Gliomas

IDH-wildtype gliomas displayed a distinct spatial profile, with half as many tumors than would be expected in the frontal lobe (O:E = 0.59; *adj-p* < 0.001), including both the SFG (O:E: = 0.57; *adj-p* = 0.03) and MFG (O:E = 0.60; *adj-p* = 0.009). Hotspots included the insula, hippocampus, superior temporal gyrus, thalamus, and temporal lobe (Table 3B). The insula had 4.6 times more tumors than expected (18 vs. 4; *adj-p* < 0.001). The temporal lobe had 1.5 more tumors than expected (50 observed vs 33 expected, *adj-p =* 0.006). Furthermore, the hippocampus showed a sixfold enrichment (12 vs. 2 expected; *adj-p* < 0.001), and the superior temporal gyrus showed a 2.1 fold enrichment (19 vs 9 expected; *adj-p* = 0.001) (Figure 2). The occipital lobe had less than half as many tumors as would be expected (O:E = 0.47; *adj-p* = 0.008), but more frequent than IDH-mutant tumors (5.5% vs 0.5%, respectively).

These patterns were largely recapitulated in the validation cohort which also showed significant enrichment in the insula (O:E = 3.75; *adj-p* = 0.001) and hippocampus (O:E = 4.5; *adj-p* = 0.01). The middle (O:E = 2.72; *adj-p* = 0.0001) and inferior (O:E = 2.63; *adj-p* = 0.003) temporal gyri had significant enrichment (see *Supplemental Digital Content*).

### Spatial Patterns by Molecular Subtype

Molecular subtype analysis showed that both IDH-mutant astrocytomas and oligodendrogliomas were enriched in the SFG (O:E = 3.72 and 3.45, respectively; both *adj-p* < 0.001) (Figure 3). The distribution within the SFG did not differ between molecular subtypes (*p* = 0.98). Astrocytomas were more likely than oligodendrogliomas to involve the insula (24 vs. 4; *adj-p* < 0.001).

**Figure 3.** Spatial comparison by molecular subtype (NYU). Both oligodendroglioma and astrocytoma were enriched in the superior frontal gyrus; only oligodendroglioma reached significance in the middle frontal gyrus. Astrocytoma showed greater insular enrichment than oligodendroglioma. * Observed counts were significantly higher than expected. ** Observed counts were significantly lower than expected.

### Genotype-Specific Regional Differences

Genotype-specific comparison confirmed the SFG predilection for IDH-mutant gliomas, which occurred 6.17 times more frequently than IDH-wildtype tumors (95% CI: 3.55-10.71) (Figure 4). In the insula, the relative risk of IDH-mutant tumor compared to IDH-wildtype was 1.54 (95% CI: 0.88-2.69), which did not reach statistical significance.

**Figure 4.** Heatmap of gliomas across discovery and validation cohort.

### Anatomical Correlates of Clinical Features

Spatial tumor location within the NYU IDH-mutant cohort did not significantly influence overall survival (p = 0.55) or progression-free survival (hazard ratio = 1.22; 95% CI: 0.70-2.12; p = 0.49) (Table 4).

**Table 4.** Patient Differences Between Gliomas Located in Superior Frontal Gyrus vs. Other Locations.

| Characteristics |  | Superior Frontal Gyrus (n=81) | Others (n=123) | p-value |
| --- | --- | --- | --- | --- |
| Sex |  |  |  | 0.65 |
|  | M | 41 | 57 |  |
|  | F | 40 | 66 |  |
| Age (Median ± SD) | | | | 0.37 |
| | | 45.7 ± 12.1 | 47.4 ± 13.9 | |
| Race |  |  |  | 0.33 |
|  | White not of Hispanic American Origin | 10 | 79 |  |
|  | Black not of Hispanic American Origin | 0 | 3 |  |
|  | Hispanic | 9 | 15 |  |
|  | Asian | 7 | 13 |  |
|  | American Indian | 2 | 2 |  |
|  | Other | 2 | 3 |  |
|  | Unknown | 13 | 8 |  |
| **Ethnicity** |  |  |  | 1.0 |
|  | Not of Spanish or Hispanic Origin | 67 | 107 |  |
|  | Hispanic or Latino or Spanish | 10 | 16 |  |
|  | Unknown | 4 | 1 |  |
| **Seizure as Initial Presentation** |  |  |  | 0.71 |
|  | Yes | 44 | 58 |  |
|  | No | 36 | 61 |  |
| **Initial Intervention** |  |  |  | 0.85 |
|  | Biopsy | 9 | 16 |  |
|  | Resection | 72 | 107 |  |
| **Second Resection Performed** |  |  |  | 0.19 |
|  | Yes | 38 | 45 |  |
|  | No | 43 | 78 |  |
| **KPS** |  |  |  | 0.12 |
|  | 100 | 22 | 22 |  |
|  | 90 | 28 | 59 |  |
|  | 80 | 11 | 16 |  |
|  | 70 | 7 | 3 |  |
|  | 60 | 4 | 4 |  |
|  | 50 | 0 | 3 |  |
|  | 40 | 0 | 1 |  |
|  | 30 | 0 | 1 |  |
| **Time to First Progression, Days (Mean)** |  |  |  | 0.12 |
|  |  | 1839 | 2514 |  |
| **Time Between First and Second Resection, Days (Mean)** |  |  |  | 0.19 |
|  |  | 1424 | 2806 |  |
| **Overall Survival** |  |  |  | 0.33 |
|  | Alive | 78 | 113 |  |
|  | Deceased | 3 | 10 |  |
| **Imaging: Enhancing at Diagnosis?** |  |  |  | 0.03 |
|  | Yes | 31 | 28 |  |
|  | No | 43 | 82 |  |
| **Laterality** |  |  |  | 1.0 |
|  | Left | 42 | 64 |  |
|  | Right | 39 | 59 |  |
| **Average Total Volume (cm3)** |  |  |  | 0.15 |
|  |  | 45.4 | 37.5 |  |
| **Pathology** |  |  |  | 0.98 |
|  | Astrocytoma | 40 | 63 |  |
|  | Oligodendroglioma | 41 | 60 |  |

### Transcriptomic Correlates of Regional Susceptibility

To investigate biological mechanisms underlying the spatial enrichment of IDH-mutant gliomas, we performed a controlled spatial gene expression study comparing (1) the frontal versus occipital lobes and (2) the superior versus inferior frontal gyri.

### Differential Expression Analysis

Between the frontal and the occipital lobes, 36 genes were differentially expressed; 28 genes were upregulated and 8 genes downregulated in the frontal lobe. No significant differential gene expression was observed between the superior and inferior frontal gyri (see *Supplemental Digital Content*).

### Gene Co-expression Network Analysis

Unsupervised clustering of co-expressed genes resulted in four modules “yellow”, “green”, “magenta”, and “black” that were statistically significant with a correlation value greater than 0.1. The “yellow” module showed an increased gene co-expression within the frontal lobe, while other modules were seen to show an increase in the occipital lobe (Figure 5). Pathways associated with each pathway are shown in Figure 4. “Yellow” module showed a weak Pearson correlation of pathways associated with *cholesterol metabolism* (NES = 1.59; *adj-p* < 0.05). No pathways were specifically enriched in the SFG (See *Supplemental Digital Content*).

**Figure 5.** Weighted gene co-expression network analysis by lobe. (A) Soft-threshold power selected at the first scale-free topology fit R^2^ ≥ 0.85 to approximate a scale-free network. (B) TOM-based hierarchical clustering with dynamic tree cut identified co-expression modules. (C) Module–lobe associations; the yellow module showed a weak positive association with the frontal lobe. (D) Volcano plot for the yellow module showing log2 fold-change versus significance. Yellow points denote module genes; red and blue indicate up- and down-regulated genes, respectively. No yellow-module genes exceeded the prespecified log2 fold-change significance threshold.

### Gene Set Enrichment Analysis

GSEA revealed several pathways enriched in the frontal versus occipital lobes, including fatty acid metabolism (NES 1.94, *adj-p* = 0.05), cholesterol homeostasis (NES 1.78, *adj-p* = 0.07), adipogenesis (NES 1.64, *adj-p* = 0.07), reactive oxygen species pathway (NES 1.62, *adj-p* = 0.08), and MTORC1 signaling (NES 1.61, *adj-p* = 0.08) (Figure 6; see *Supplemental Digital Content*). Cholesterol metabolism was the only pathway consistently identified in both GSEA and WGCNA. Pathway PID_P53_Regulation_Pathway was enriched in the superior frontal gyrus (NES 1.75, *adj p-val* = 0.23).

**Figure 6.** Differential expression and pathway enrichment between frontal and occipital cortex. (A) Volcano plot of frontal vs. occipital differential gene expression. Significance for enrichment was set at absolute log-fold change ≥ 1 and adjusted p-value < 0.05, which revealed 28 upregulated and 8 downregulated genes in the frontal lobe. GSEA analysis was then performed on the ranked gene list (B) and also on a restricted gene list from the WGCNA yellow module (C).

## Discussion

This multi-institutional analysis of 821 gliomas reveals a striking anatomical bias in the distribution of IDH-mutant tumors, with a disproportionate enrichment in the SFG. Over one third (35.6%) of IDH-mutant gliomas in the primary cohort arose in the SFG - over four times more frequently than IDH-wildtype tumors. Notably, IDH-mutant gliomas showed a clear midline preference, evident by the observed to expected ratio the superior frontal gyrus (O:E = 3.5), which gradually tapers laterally with the middle frontal gyrus (O:E = 2.2) before reaching the inferior frontal gyrus (O:E = 1) in a stepwise fashion. This regional predilection was shared by both IDH-mutant astrocytomas and oligodendrogliomas, suggesting that the spatial signal reflects IDH mutation status rather than histologic subtype. In contrast, IDH-wildtype gliomas demonstrated preferential localization to limbic and sensorimotor cortices, including hippocampus and temporal cortex, underscoring a reproducible genotype-specific vulnerability. While previous studies have noted frontal lobe enrichment of IDH-mutant gliomas, they often treated the entire frontal lobe as a single structure and lacked further investigation into individual gyri and molecular stratification.^10,22,23,28^ Some studies pooled IDH-mutant and wildtype tumors, limiting the ability to assess genotype-specific spatial differences.^21^ Our atlas-based approach enabled high-resolution mapping, refining the epicenter of IDH-mutant glioma origin to the SFG and delineating an anterior–posterior gradient. Conversely, the limbic and temporal predilection of IDH-wildtype gliomas aligns with prior observations in high-grade glioma cohorts.^29,30^

Despite robust spatial differences, transcriptomic analyses revealed only modest molecular correlates. Differential expression between the frontal and occipital lobes identified 36 genes, but no significant pathways from GSEA analysis overlapped with these genes. This is consistent with prior observations that transcriptomic variation across cortical gyri is often subtle.^31^ These findings suggest that large-amplitude changes in single gene expression are unlikely to explain the anatomical predilection of IDH mutant gliomas.

Co-expression network and gene set enrichment analyses yielded modest but convergent signal implicating metabolic pathways Specifically, the “yellow” WGCNA module, enriched in the frontal lobe, was associated with cholesterol and fatty acid metabolism – pathways also identified in GSEA comparisons between frontal and occipital regions. However, many constituent genes did not meet fold-change thresholds for differential expression, indicating that region-specific transcriptomic differences are small yet coordinated, possibly demonstrating subtle, multicentric shifts in expression that may reflect microenvironmental or cell type compositional differences rather than large amplitude changes in individual transcripts.

These findings raise the possibility that the frontal lobe, and the SFG in particular, harbors a distinct cellular and metabolic microenvironment conducive to IDH-mutant gliomagenesis. Astrocytes and oligodendrocytes play a crucial role in cholesterol transportation and synthesis in the central nervous system (CNS), where the blood-brain barrier restricts plasma lipids entering the CNS.^32,33^ Brain cholesterol is vital for membrane stability, synaptogenesis, and saltatory conduction in the CNS.^34–36^ IDH-1 mutations deplete NADPH and disrupt lipid homeostasis, thereby disrupting these essential neurobiological processes.^37–39^ Mouse models of IDH-1 mutant glioma have demonstrated regional cholesterol depletion and structural disruption in frontal white matter, suggesting that focal cholesterol dysregulation may establish a permissive microenvironment via altered membrane composition or sustained glial inflammatory signaling for IDH mutant gliomagenesis.^40^ We hypothesize that the intersection of metabolic demand and IDH-mediated disruption may render the frontal cortex, and particularly the SFG, uniquely vulnerable to tumor initiation.

An additional explanation lies in the developmental lineage. Single-cell transcriptomic and epigenomic profiling has demonstrated that the SFG and adjacent association cortices are enriched for oligodendrocyte progenitor-like and neural precursor-like cells, which are implicated as the cells of origin for IDH-mutant gliomas.^41–46^ These progenitor populations exhibit chromatin states and gene expression profiles that are permissive to transformation by IDH mutation, including susceptibility to IDH driven epigenetic reprogramming that impairs normal differentiation and promotes tumorigenesis.^46^

Circuit-level factors may also contribute. The SFG participates in high-order association networks characterised by dense connectivity and persistent synaptic activity.^47–50^ These features may provide trophic support for early mutant cell survival, although this remains speculative in the absence of direct in vivo evidence.

Clinically, recognition of the SFG predilection carries diagnostic and therapeutic implications. A non-enhancing lesion in the SFG of a young adult should heighten suspicion for IDH-mutant pathology and may influence preoperative counseling, biopsy targeting and molecular testing algorithms. SFG involvement was not associated with inferior survival, supporting the role of maximal-safe, navigation-guided resection when feasible. Incorporating anatomic sites into prognostic models may further refine surveillance and therapeutic strategies.

This study has limitations. Its retrospective design introduces potential referral and imaging biases. Manual tumor segmentation and atlas registration carry intrinsic spatial error on the order of 1mm.^51^ Expected tumor frequencies were derived from voxel-based regional volume rather than true cellular density. Clinical covariates were incomplete for some cases. The transcriptomic analyses relied on a public dataset with six neurotypical brains. Larger, prospective studies integrating imaging, multi omic profiling, and spatially resolved epigenomic and transcriptomic analyses, will be necessary to validate these findings. Future directions include incorporating longitudinal glioma progression datasets, single-cell epigenomics, and connectomic data to better understand region-specific vulnerabilities.

In summary, we provide the first multi-institutional, high-resolution, genotype-stratified atlas of diffuse glioma localization, demonstrating that IDH-mutant gliomas exhibit a reproducible and disproportionate predilection for the superior frontal gyrus. This spatial dichotomy between IDH-mutant and IDH-wildtype tumors may reflect underlying metabolic, developmental, and circuit-level differences in regional brain biology. Deciphering the region-specific vulnerabilities that underlie this spatial predilection may illuminate fundamental principles of glioma initiation - linking developmental lineage, microenvironmental context, and molecular susceptibility in the earliest stages of tumor formation.

## Supporting information

Supplemental Digital Material

## Data Availability

All data produced in the present study are available upon reasonable request to the authors

---

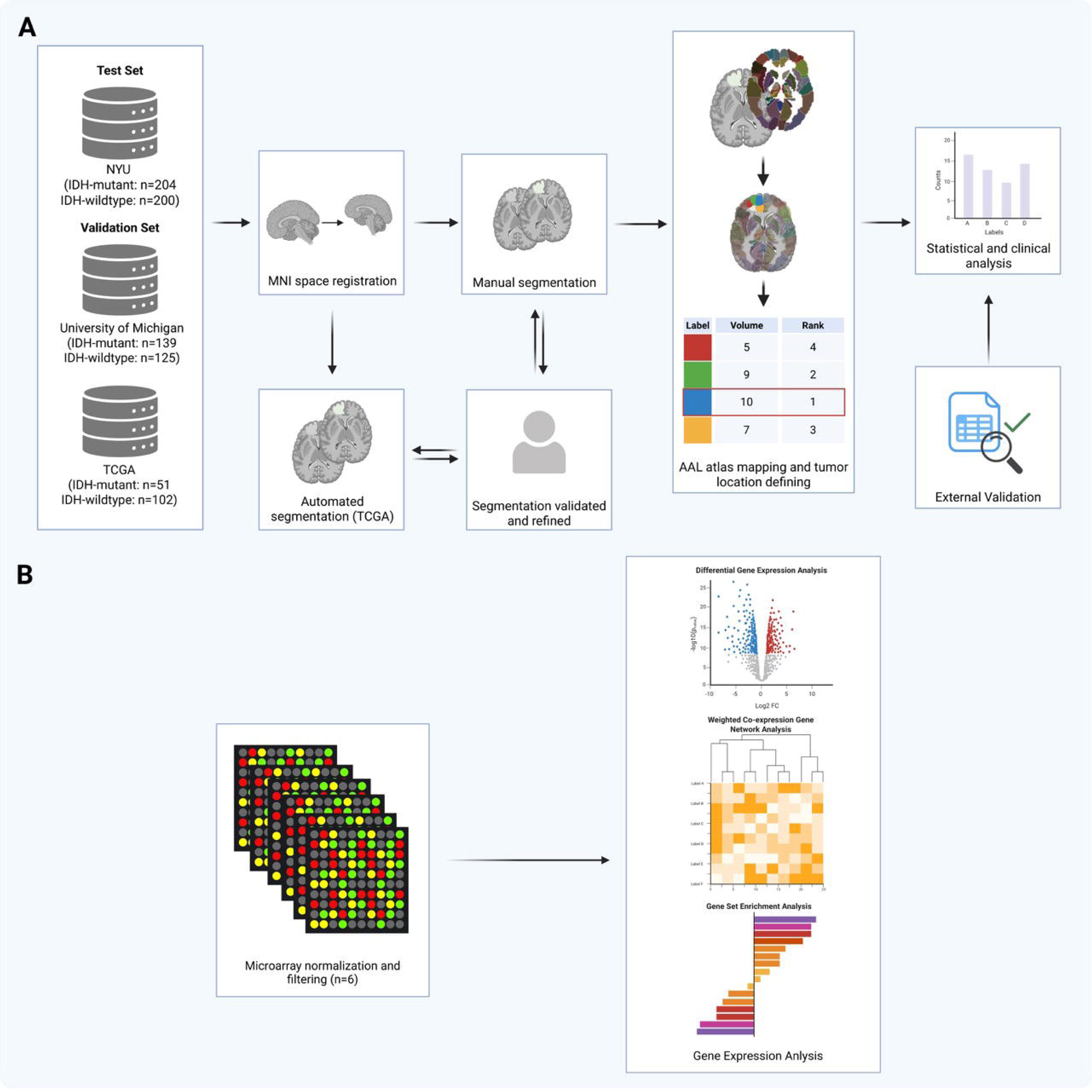

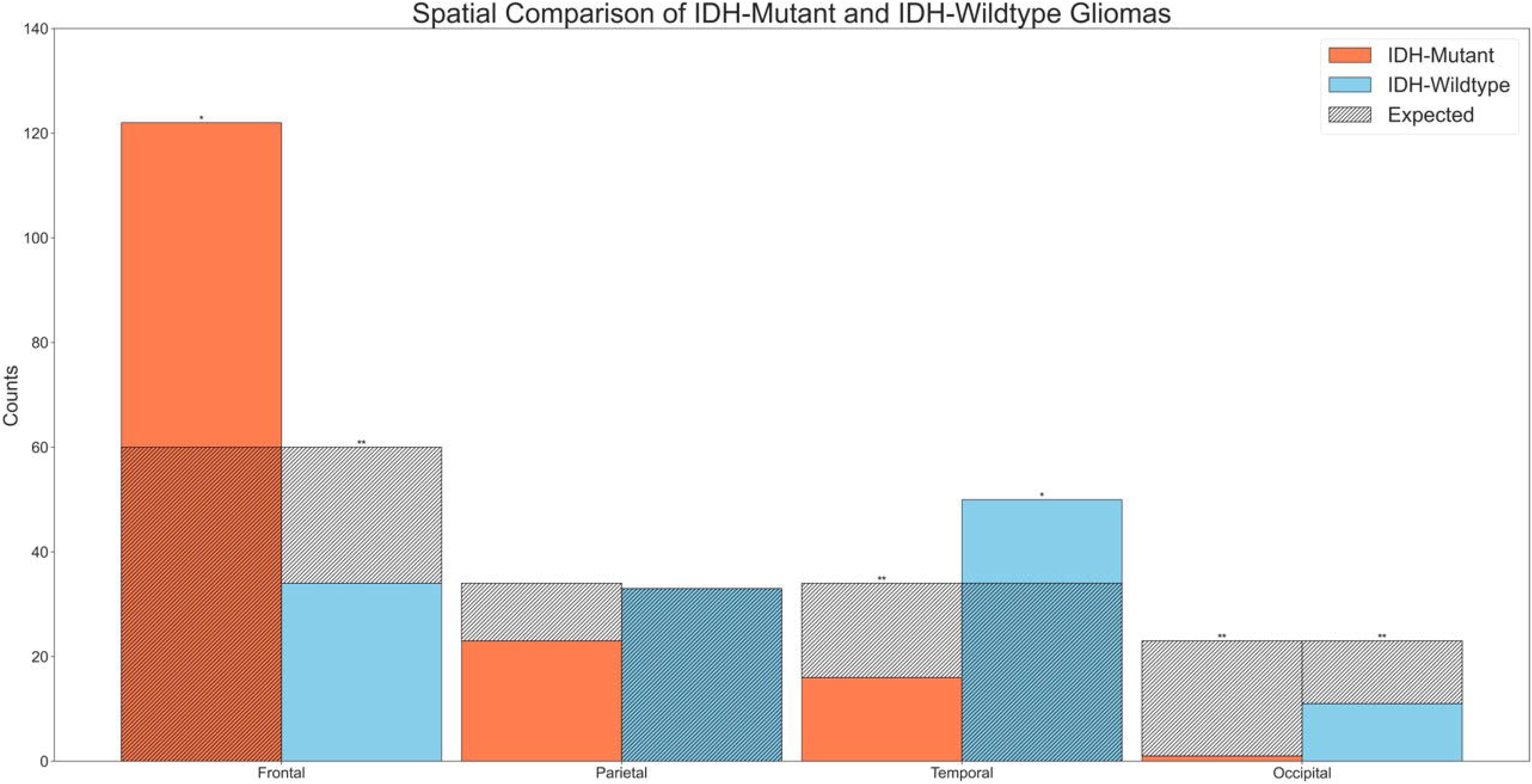

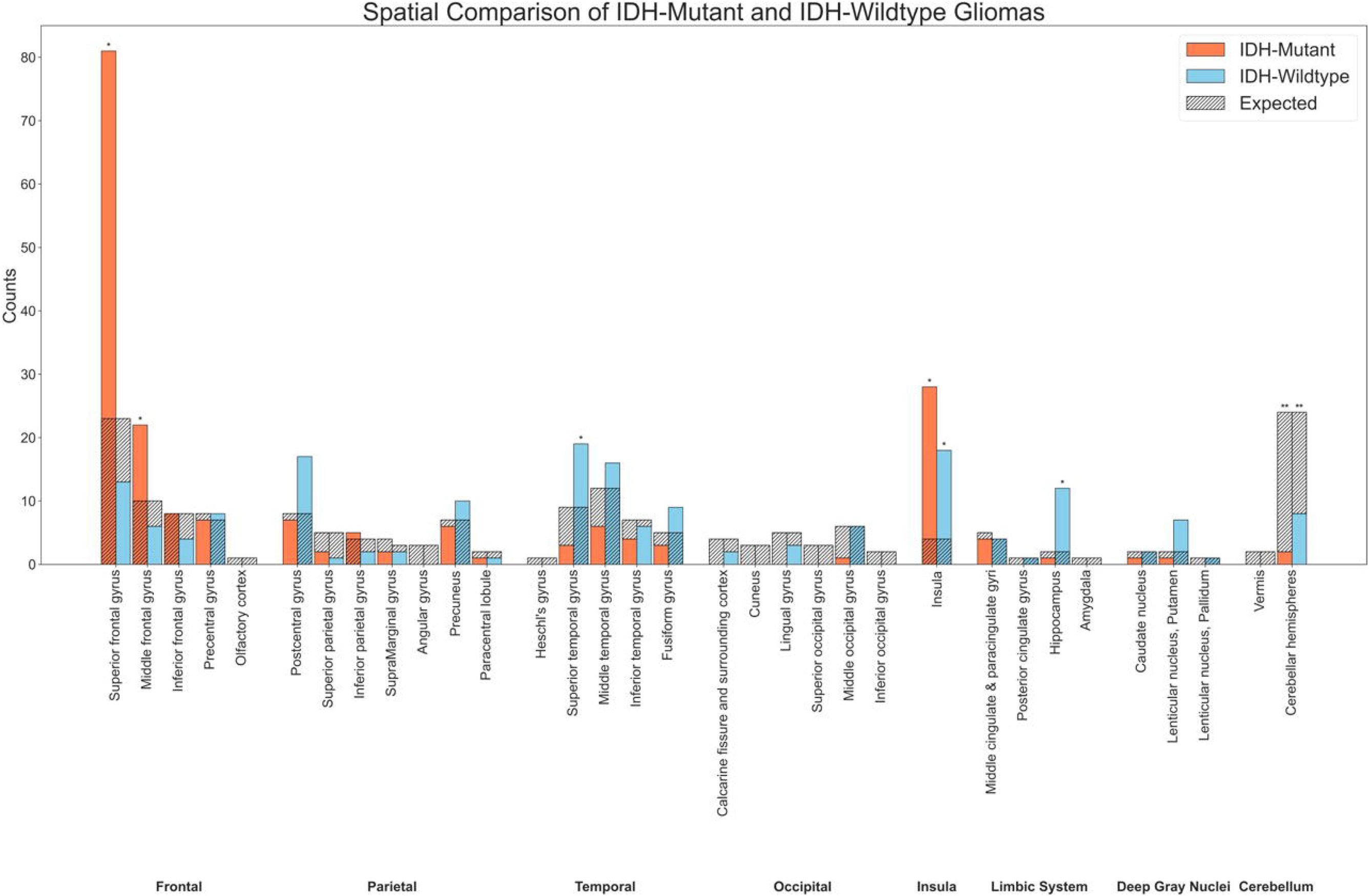

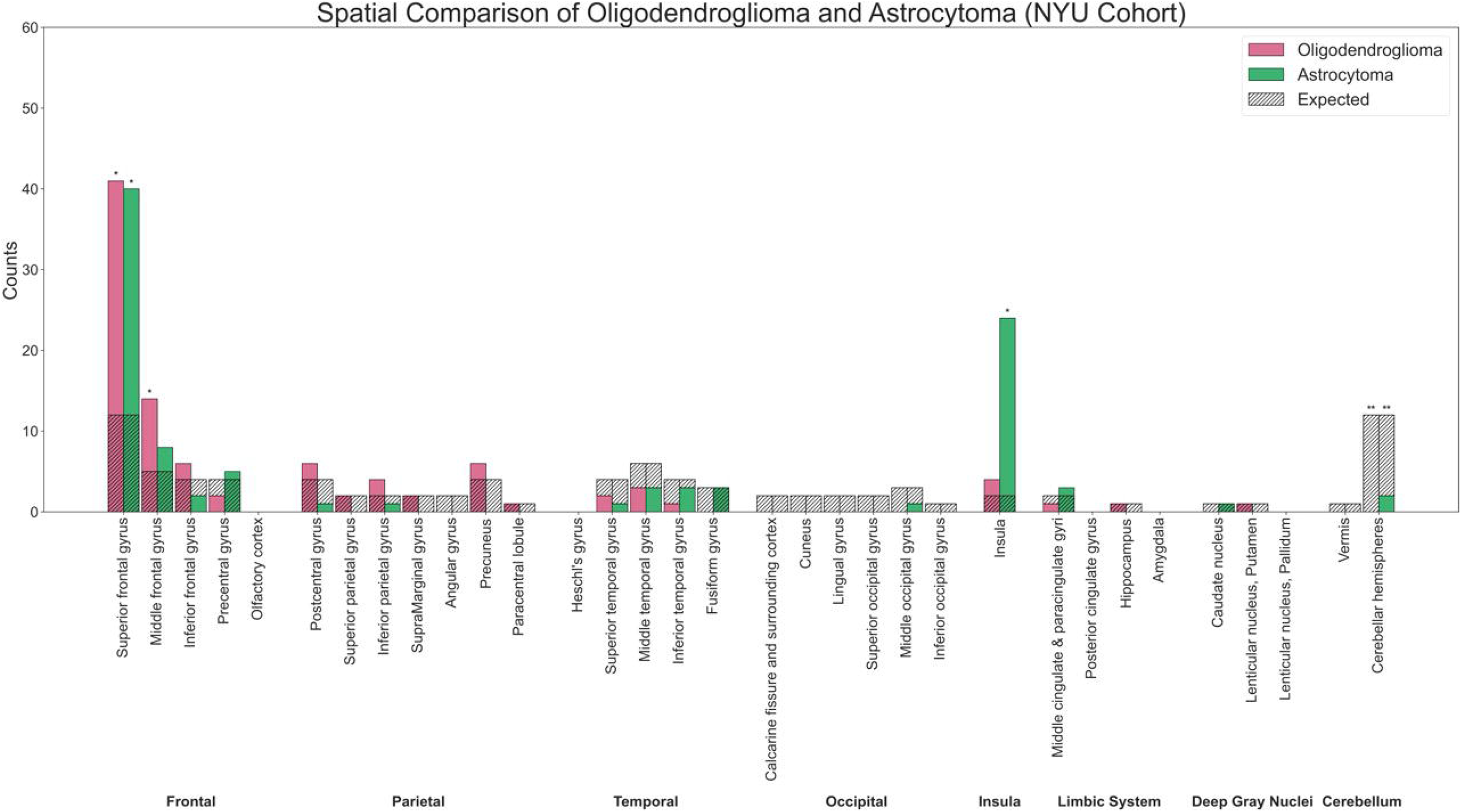

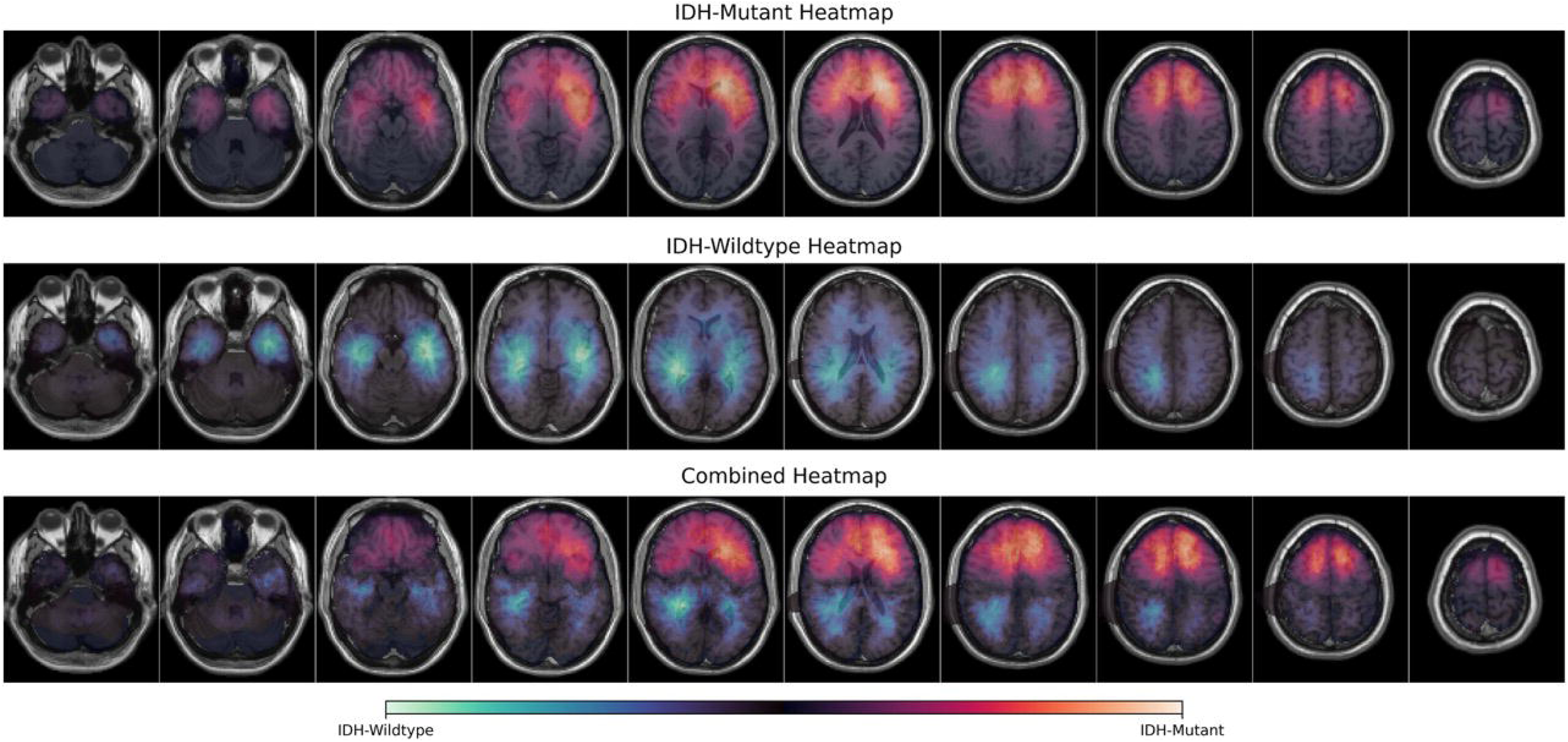

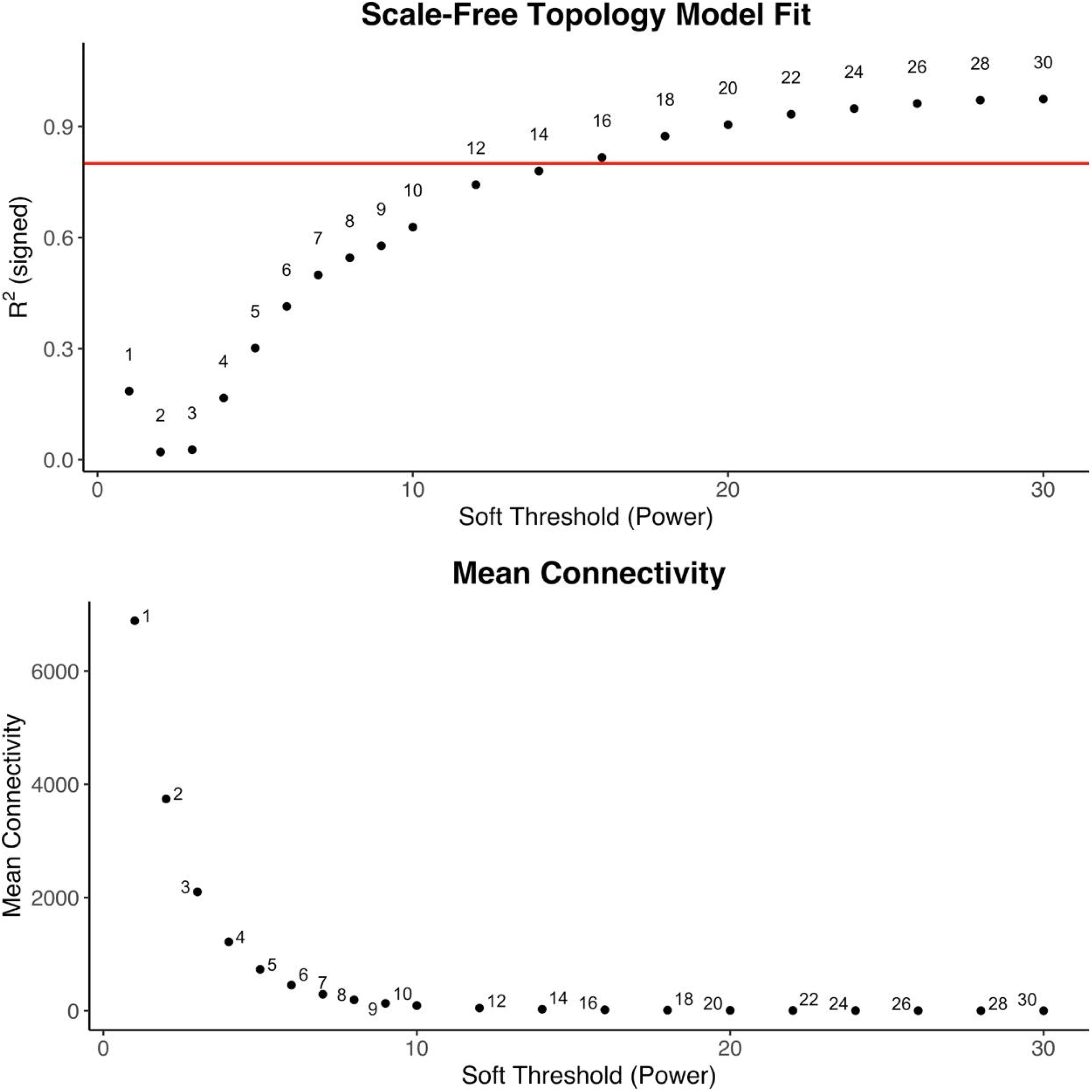

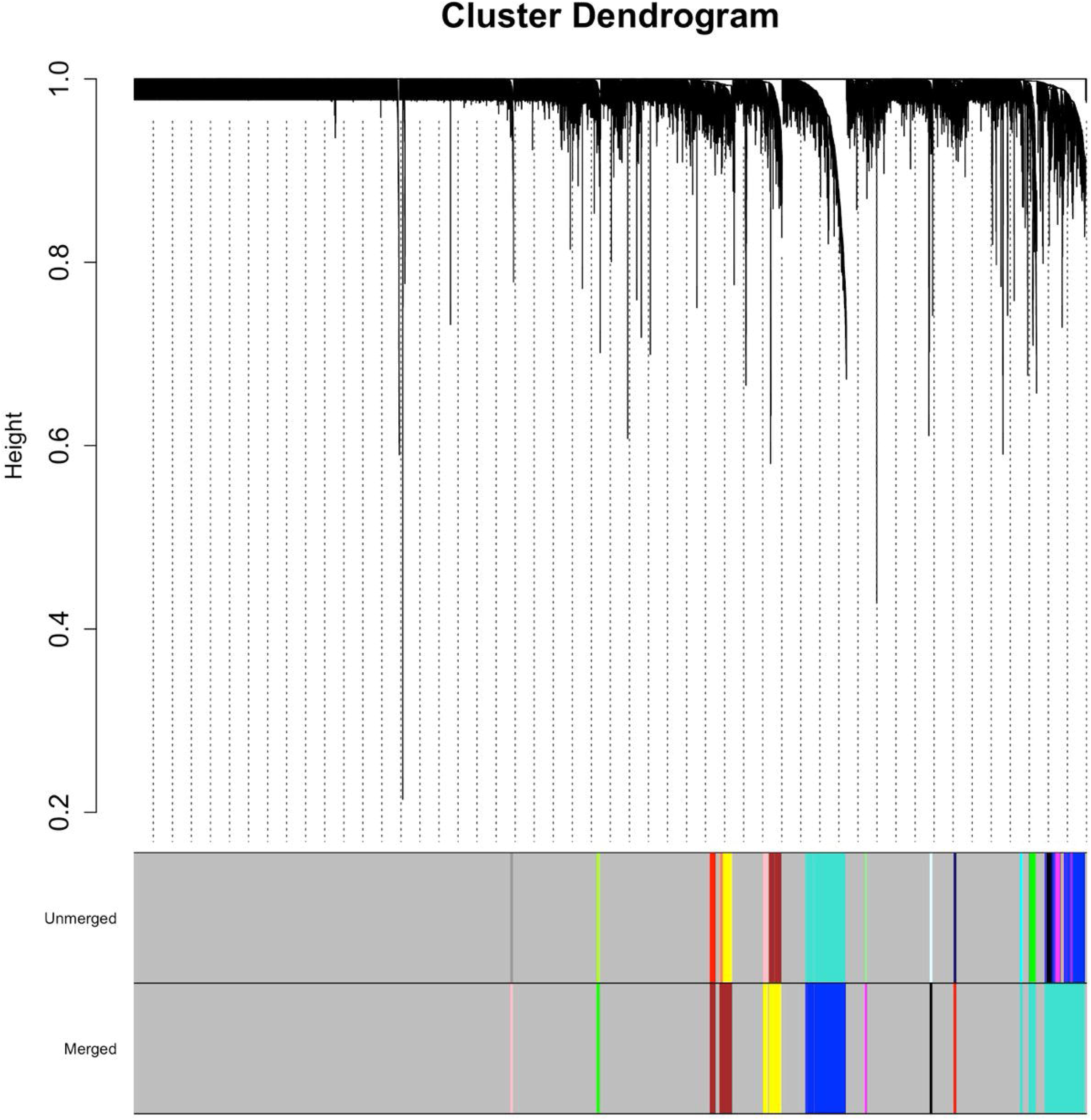

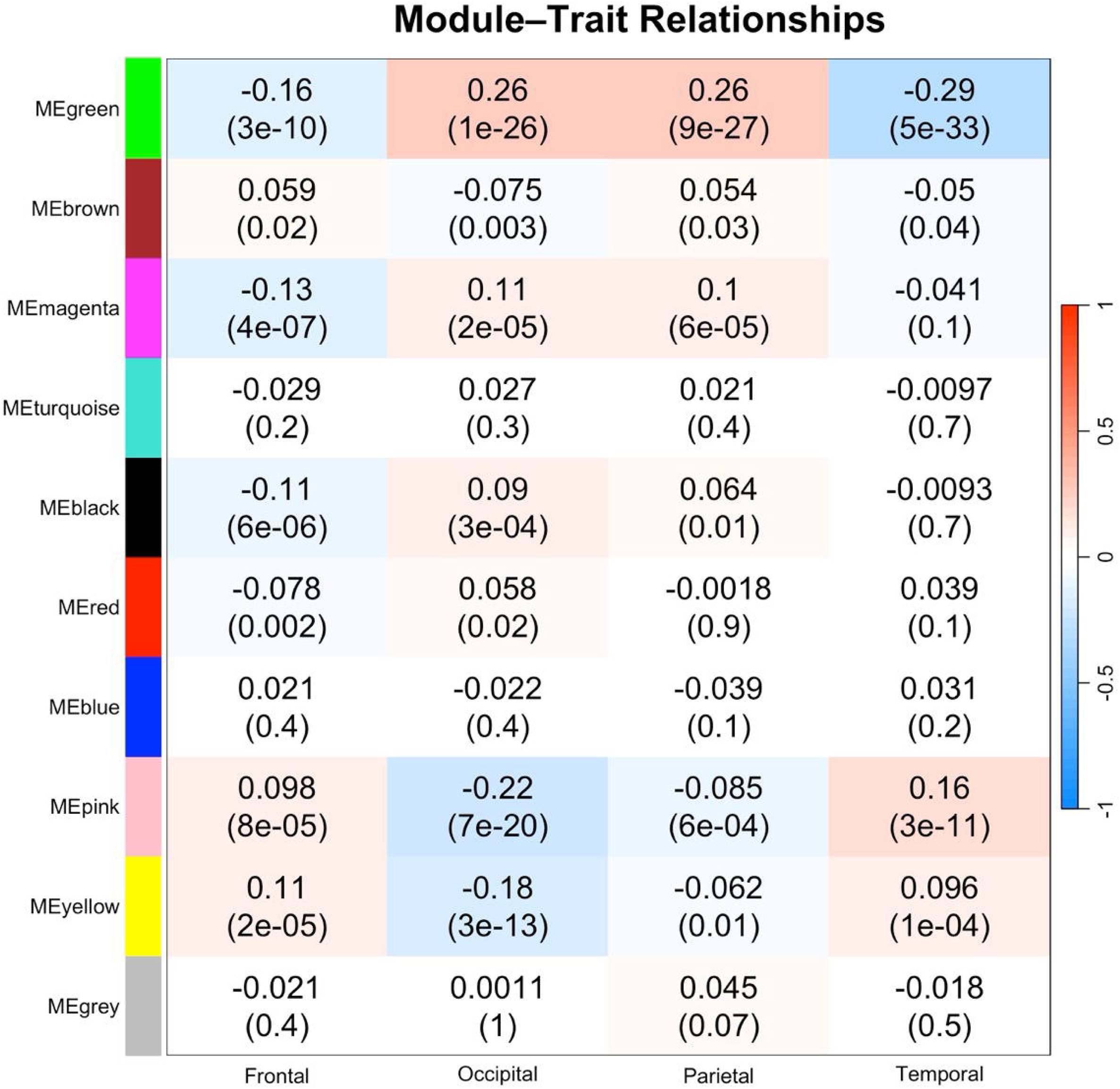

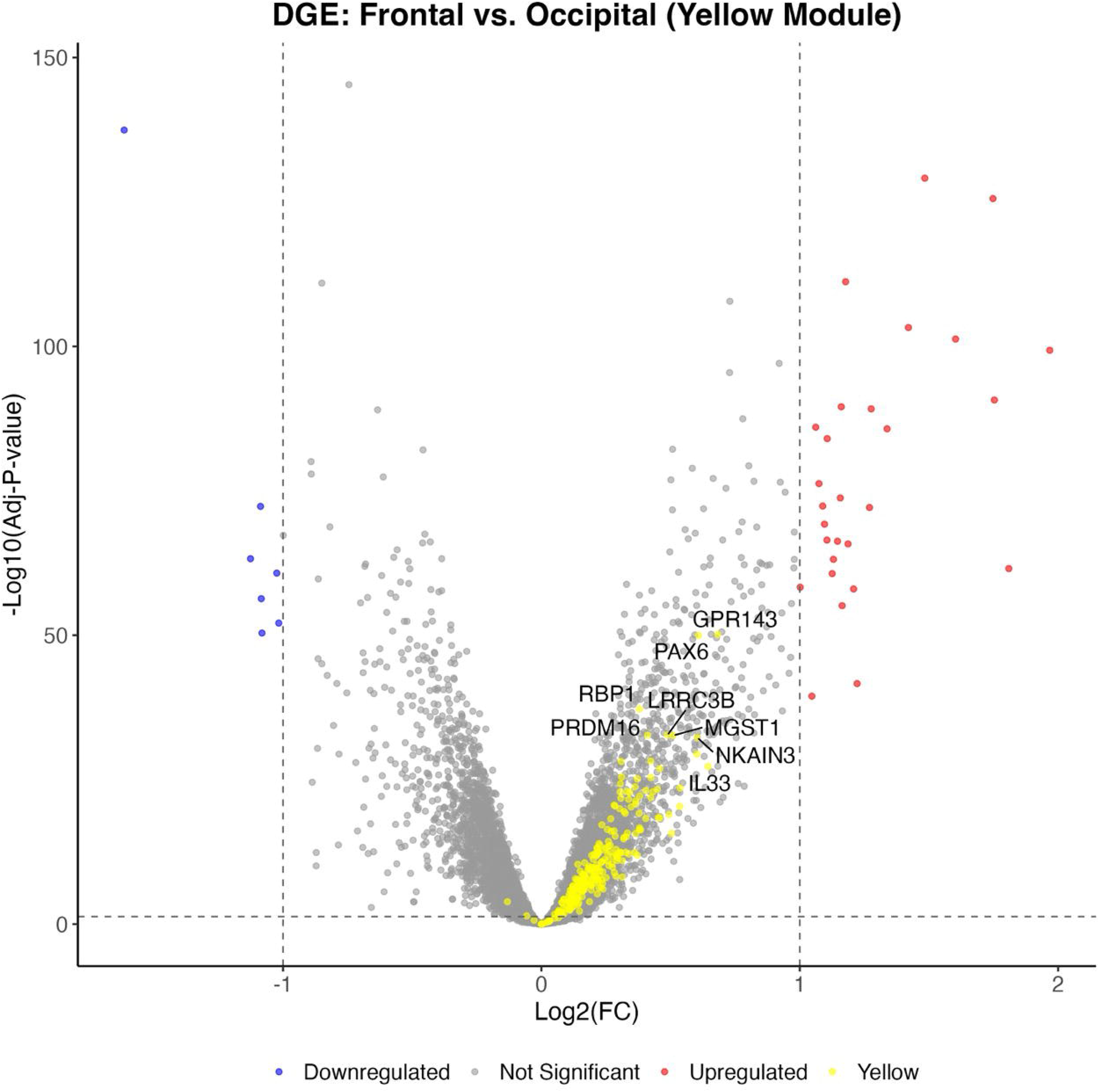

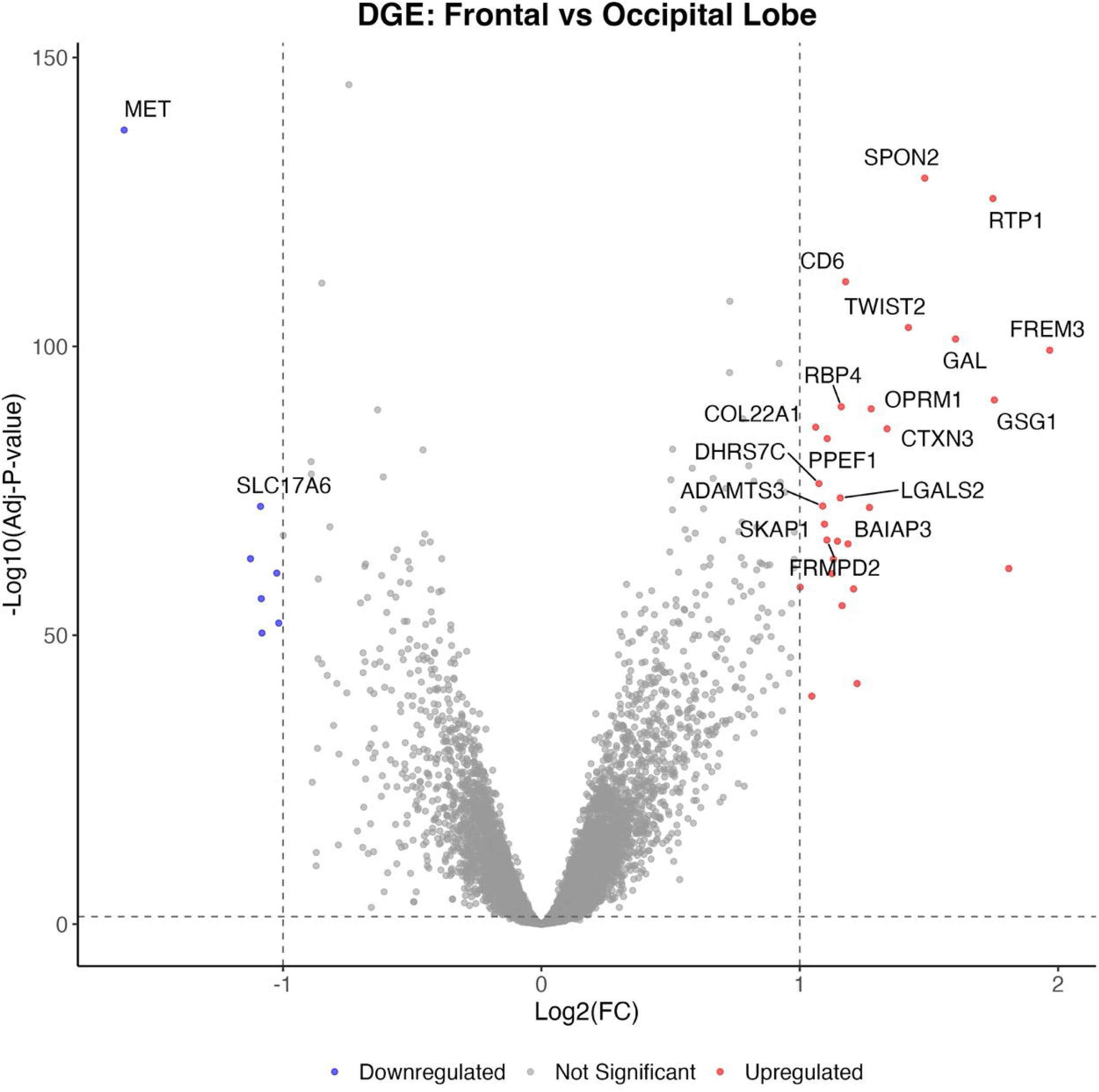

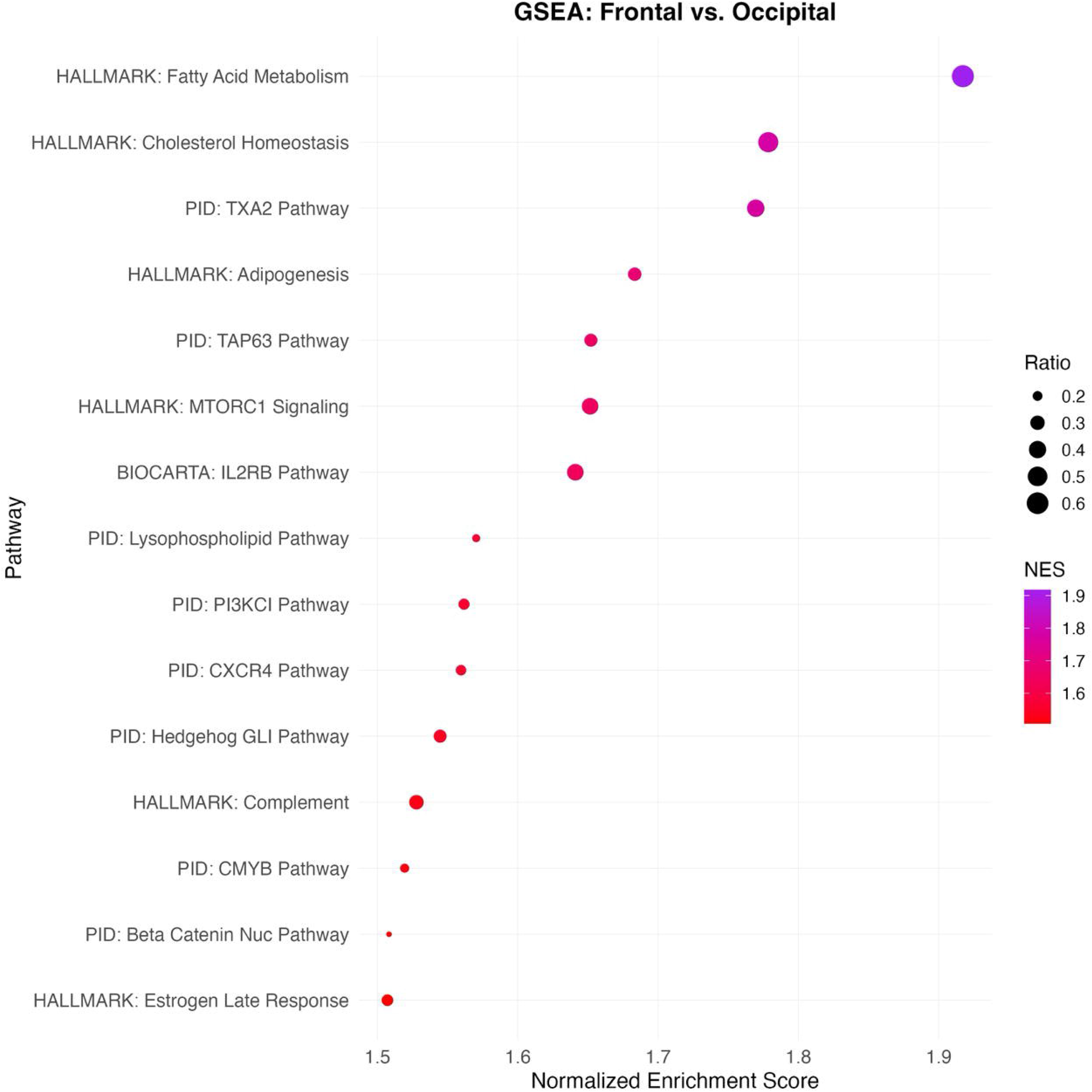

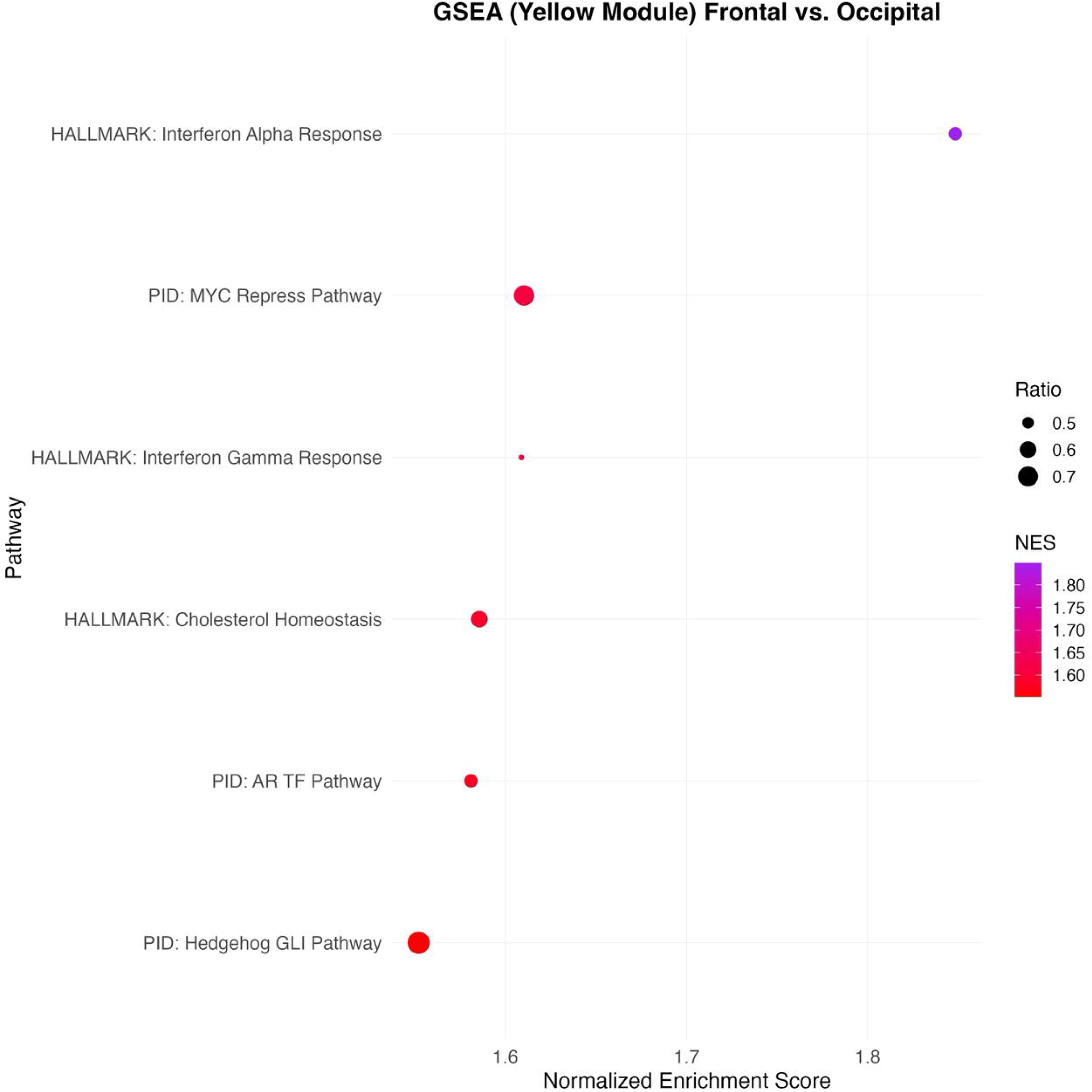

