## Supplemental Digital Material for "Anatomic Predilection of IDH-Mutant Gliomas: A Multi-Institutional Spatial Analysis"

**Supplemental Methods**

*Data collection, Image Registration, Tumor Segmentation, Location Mapping*

All magnetic resonance imaging (MRI) scans were pre-processed with Statistical Parametric Mapping 12 (SPM12), applying affine, linear, and nonlinear registration to MNI space. Each co-registered dataset, including T1, contrast-enhanced T1, T2, and FLAIR sequences, was imported into 3D Slicer (version 4.11.0) for manual tumor segmentation. Pre-segmented files were utilized for the TCGA dataset, and edema was removed as a region of interest within these segmentation files. Data processing was further done in Python. All segmentations were independently reviewed by a second investigator and iteratively refined until full consensus was reached. Two IDH-mutant tumors from the NYU cohort were subsequently excluded from volumetric analysis due to indistinct margins that precluded consensus.

Regarding locational mapping, automated anatomical labeling atlas (AAL3) was chosen due to its robust MNI‐aligned parcellation of both cortical and subcortical structures. Each anatomical atlas was initially reviewed by a neurosurgeon and certain anatomical labels were grouped into one label for clinical interpretation. The following parcellized areas were added into individual groups after atlas review:

- “Superior frontal gyrus”: 'Frontal_Sup_2', 'Frontal_Sup_Medial', 'Frontal_Med_Orb', 'Supp_Motor_Area'
- “Inferior frontal gyrus”: 'Frontal_Inf_Tri', 'Frontal_Inf_Oper', 'Frontal_Inf_Orb'
- “Anterior cingulate cortex”: 'ACC_sub', 'ACC_pre','ACC_sup'
- “Cerebellar hemispheres”: 'Cerebellum_Crus1', 'Cerebellum_Crus2', 'Cerebellum_3', 'Cerebellum_4_5’, 'Cerebellum_6', 'Cerebellum_7b', 'Cerebellum_8', 'Cerebellum_9', 'Cerebellum_10'
- “Vermis”: 'Vermis_1_2', 'Vermis_3', 'Vermis_4_5', 'Vermis_6', 'Vermis_7', 'Vermis_8', 'Vermis_9', 'Vermis_10'
- “Superior temporal gyrus”: 'Superior temporal gyrus','Temporal pole: superior temporal gyrus'
- “Middle temporal gyrus”: 'Middle tepmoral gyrus','Temporal pole: middle temporal gyrus'

*Gene Expression Analysis Data Preparation*

Six microarray data were normalized as the following in R (version 4.5.1). First, each sample was mapped to a parcellated area in the AAL3 atlas based on the MNI coordinates provided by the dataset. Second, to ensure cortical variances were detected rather than structural differences among subcortical and cerebellar areas, microarray samples that were mapped to non cortical structures were removed. Probes without Entrez ID associated were removed. Then, samples exhibiting low inter‑array correlation and probes detected in fewer than 1% of samples were removed. Probes that code for non-protein coding genes such as pseudogenes or long non-coding RNAs were removed. Then, to select the best probe among probes that map to the same gene, WGCNA’s *collapseRows* algorithm was utilized to select the probe with the max variance. The remaining probes were selected to be utilized for all six brains. Outliers were also removed using principal component analysis; any sample with an absolute Z-score greater than 2 on either PC axis was flagged as an outlier and removed.

Batch effect was observed within the dataset, with one brain versus the others. This effect was corrected through ComBat, and resulting PCA analysis revealed the batch effect to be resolved. In order to ensure that batch effect fix did not affect downstream analysis, all data analysis mentioned in the manuscript and supplemental digital content was done excluding the brain with notable batch effect, and results remained similar. Thus, we decided to include the dataset that has been adjusted for batch effect to be utilized in our downstream analysis.

For gene expression analysis, weighted gene co-expression network analysis (WGCNA), differential gene expression analysis, and gene set enrichment analysis (GSEA) was performed. Briefly, WGCNA constructs a network of gene co-expression to detect modules of highly correlated genes, which can then be compared with different regions to see if de novo gene-modules are associated with a region. Differential gene expression analysis pinpoints genes whose expression levels differ significantly between conditions or groups, helping to identify molecular drivers of the phenotype. Lastly, GSEA tests whether predefined gene sets, such as pathways or functional categories, show coordinated up- or down-regulation between groups, providing a pathway-level view of the data.

*Weighted Gene Co-Expression Network Generation (WGCNA)*

To investigate spatially discrete co‑expression patterns, we constructed a weighted gene co‑expression network using the WGCNA package in R.^29^ The soft‑thresholding power was selected at the point at which the scale‑free topology fit index first exceeded 0.80 to generate a scale-free network. Minimum module size was set to 30. Modules exhibiting statistically significant association (p < 0.05) were then further examined through preranked gene set enrichment analysis, ranking genes by module membership to observe biological pathways enriched within each module.

Normalized expression data were analyzed using the limma package. A linear model was fitted for each gene with the brain specimen batch included as a blocking factor. Empirical Bayes moderation of the standard errors was applied to enhance statistical power. Genes with absolute log_2_ fold change greater than 1 and Benjamini–Hochberg adjusted p-value less than 0.05 were defined as differentially expressed.

We performed GSEA to detect pathway‑level differences between regions of interest and modules of interest revealed from WGCNA analysis. Furthermore, GSEA was performed on the entire dataset with phenotype differences set as either 1) frontal versus occipital lobe or 2) superior frontal gyrus or inferior frontal gyrus. In order to increase statistical power, the total number of gene sets to be tested in the GSEA run was reduced as much as possible. A combination of Hallmark, BioCarta, and PID gene sets were chosen from MSigDB, retaining gene sets with size between 30 to 300 genes to improve statistical power. Phenotype shuffling was performed and the number of permutations was set to 1000. For standard GSEA runs, pathways were considered to be significant at a nominal P < 0.05 and FDR < 0.25 per GSEA guidelines. For pre-ranked GSEA runs, nominal p-values < 0.05 and FDR < 0.05 were used to state significance.

*Statistical Analysis*

Descriptive analyses were conducted using Python (v3.12.7) with the pandas (v2.2.3), NumPy (v2.0.1), and SciPy (v1.15.2) libraries. Figures were generated using Matplotlib (v3.10.1) and Seaborn (v0.13.2).

The expected distribution of tumors across anatomical regions was estimated by dividing the voxel count of each atlas label by the total voxel count in the atlas. The expected number of tumors per region was then calculated by multiplying this ratio by the cohort size, under the assumption of uniform voxel-based probability of tumor origin. For anatomical data analysis, p-values were adjusted for multiple comparisons using Benjamini-Hochberg false discovery rate method, and adjusted p-values < 0.05 were considered statistically significant. Relative risk of tumor localization was determined with IDH mutation status as the exposure and tumor assignment to predefined regions of interest as the outcome.

**Results**

*Spatial Tumor Analysis*

For the IDH-mutant validation cohort, a total of 59 (31.1%) tumors were found in the superior frontal gyrus, a 2.7 fold increase from expected (adj p-val < 0.001). 104 (54.7%) tumors arose in the frontal lobe, while only 2 (1.1%) tumors rose from the occipital lobe. Superior frontal gyrus (adj p-val < 0.001), middle frontal gyrus (adj p-val = 0.02), and the insula (adj p-val < 0.001) were all noted to be significantly elevated when compared to the expected tumor count by gyral volume (Supplemental Figure 1).

The IDH-wildtype validation cohort showed similar results in that it showed an increase in the temporal lobe with a 1.92 fold increase compared to expected (observed vs expected, 73 vs 38; adj p-val < 0.001). The hippocampus (9 vs 2; adj p-val = 0.01) and the insula (15 vs 4; adj p-val = 0.002) were also shown to be elevated, reflecting the findings in the NYU cohort. The validation cohort did show an increase of tumors in the middle temporal (30 vs 11; adj p-val = 0.0001) and inferior temporal gyrus (21 vs 8; adj p-val = 0.003), which was not seen in the NYU cohort (Supplemental Figure 2).

*Gene Expression*

The superior frontal gyrus did not show any differentially expressed genes when the log fold change cut off was set to 1. WGCNA analysis revealed no pathways that were elevated within the superior frontal gyrus. However, there were two modules that were elevated in the anterior cingulate cortex and the precentral gyrus (Supplemental Figure 3). The “red” module, which was associated with the precentral gyrus (r = 0.5, p < 0.001) reported one significant pathway HALLMARK_OXIDATIVE_PHOSPHOYRLATION (NES 1.89, adj p-val < 0.001). The “brown” module, which was associated with the anterior cingulate cortex (r = 0.32, p < 0.001), reported pathways involving inflammatory and cholesterol metabolism pathways (Supplemental Figure 4).

**Supplemental Figure 1.** IDH-Mutant Glioma Distribution Between NYU and Validation Cohort

**Supplemental Figure 2.** IDH-Wildtype Glioma Distribution Between NYU and Validation Cohort

**Supplemental Figure 3.** Module-trait Relationships within the Frontal Lobe

**Supplemental Figure 4.** Gene Set Analysis on Brown Module
