## Supplementary figures and images for "Anatomic Predilection of IDH-Mutant Gliomas: A Multi-Institutional Spatial Analysis"

### Supplemental Figure 1A

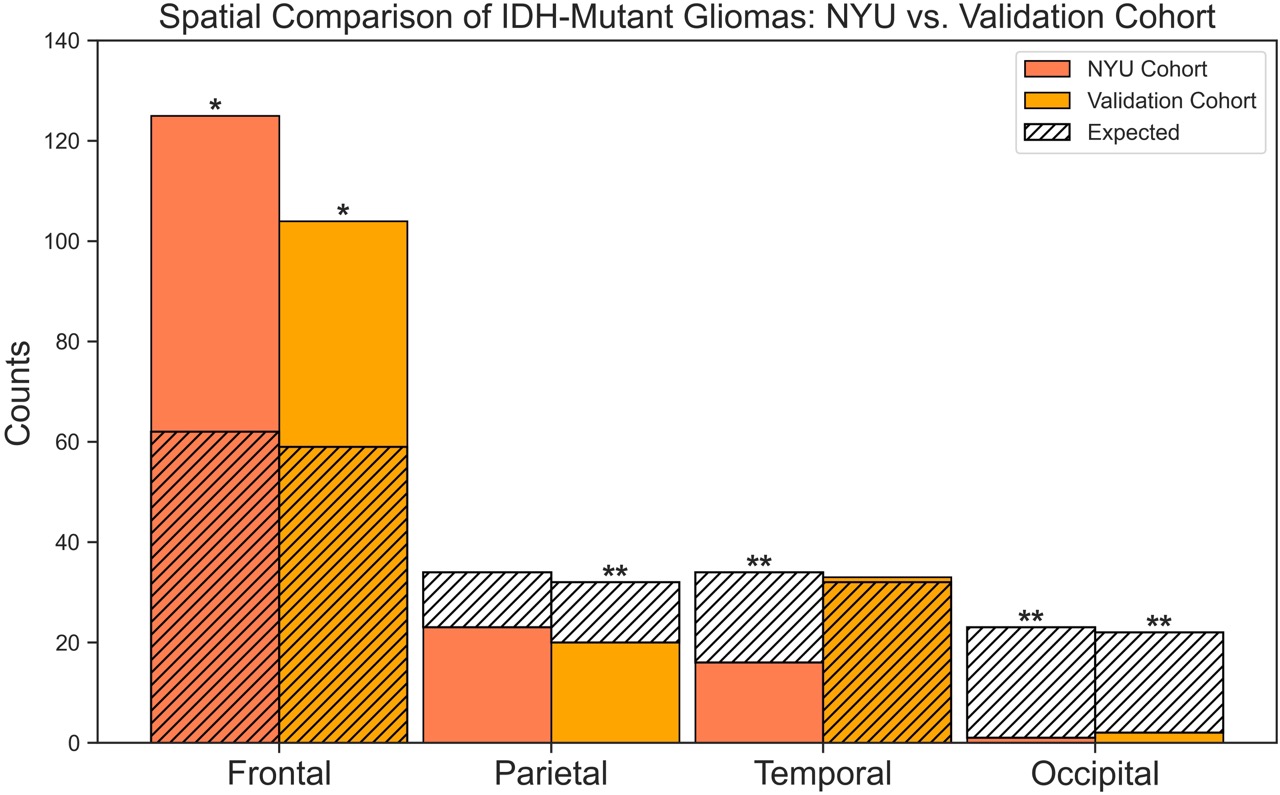

### Supplemental Figure 1B

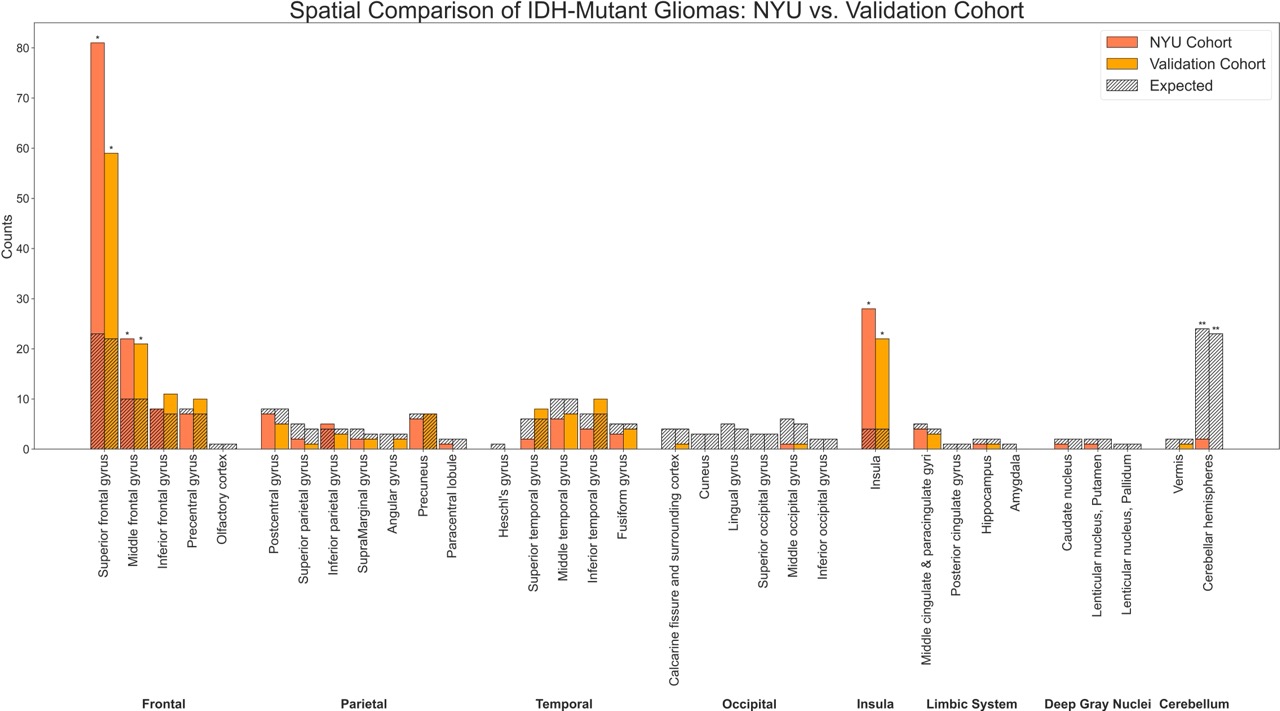

### Supplemental Figure 2A

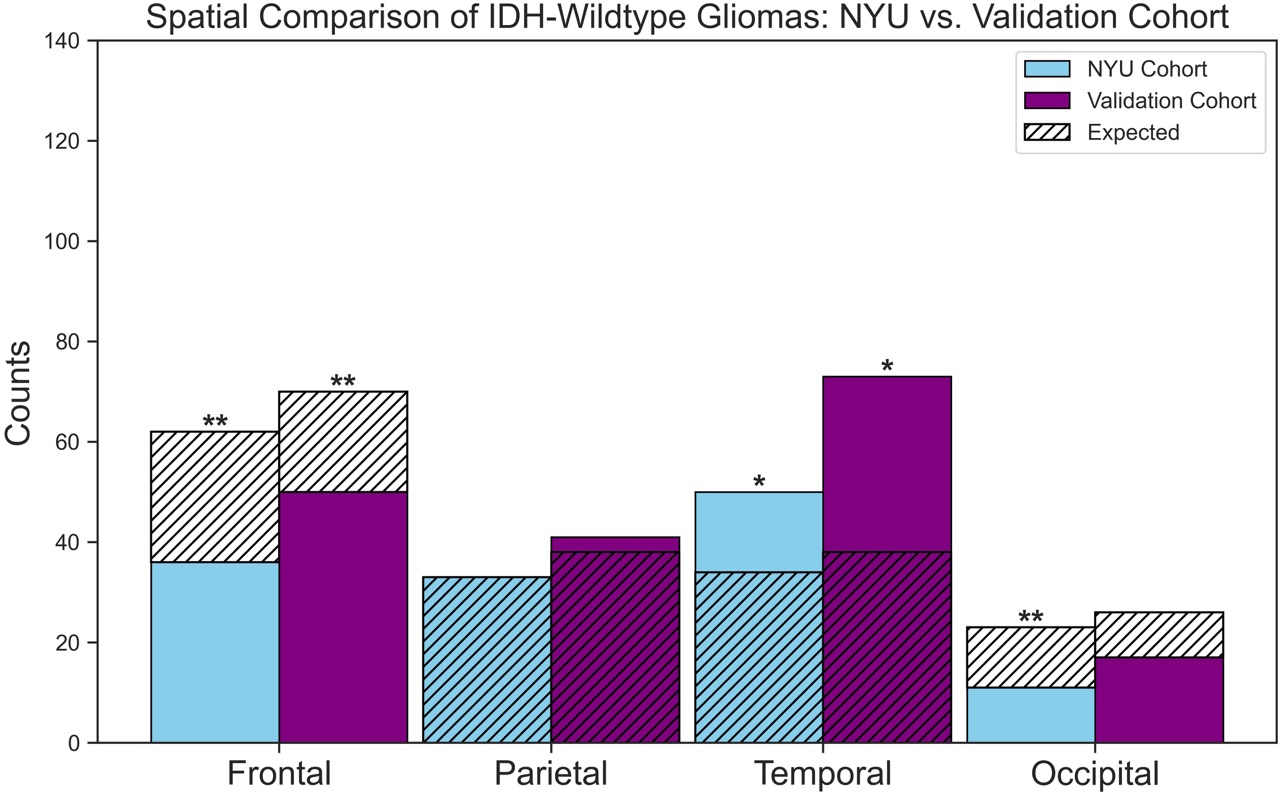

### Supplemental Figure 2B

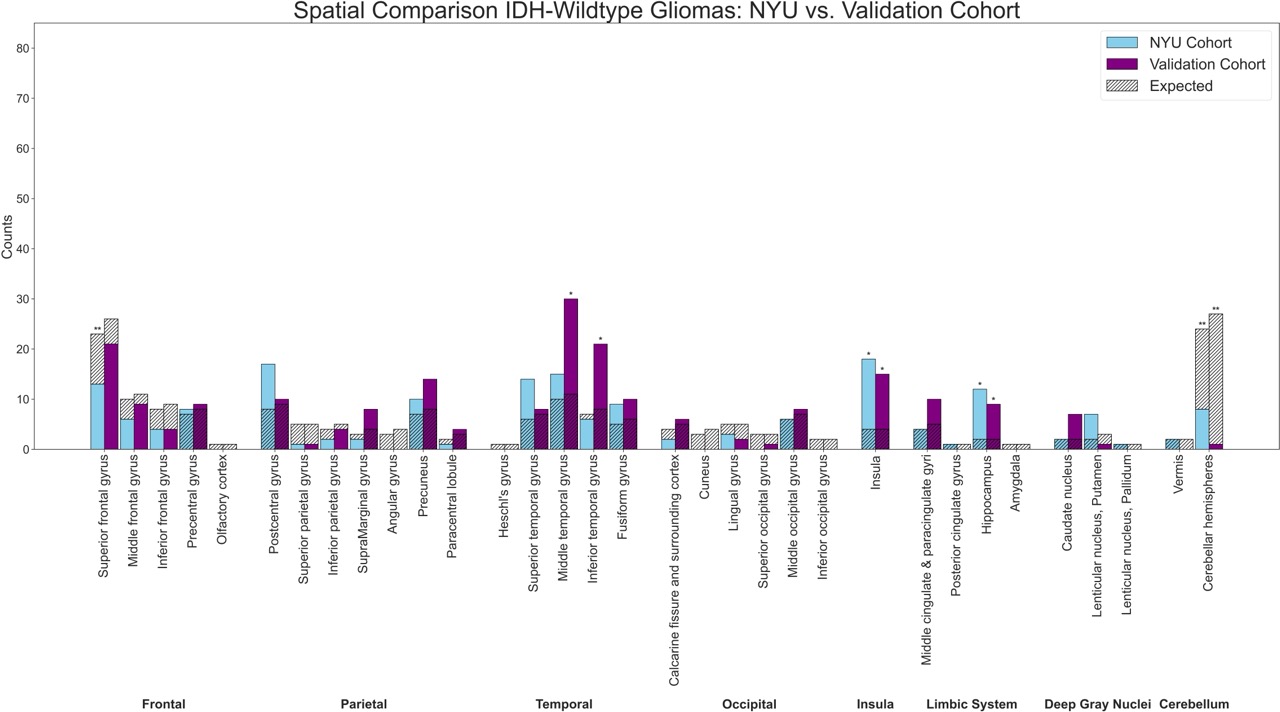

### Supplemental Figure 3

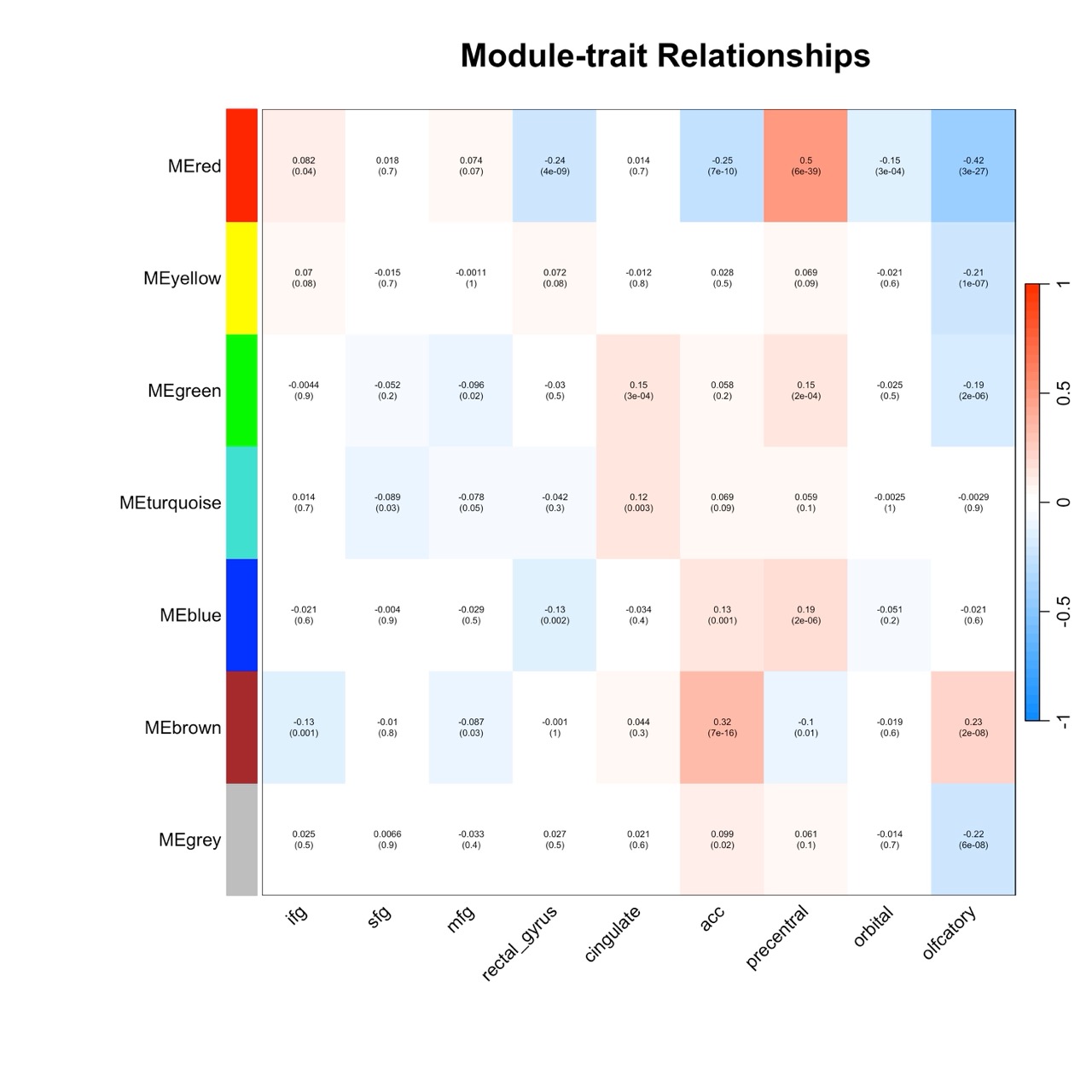

### Supplemental Figure 4

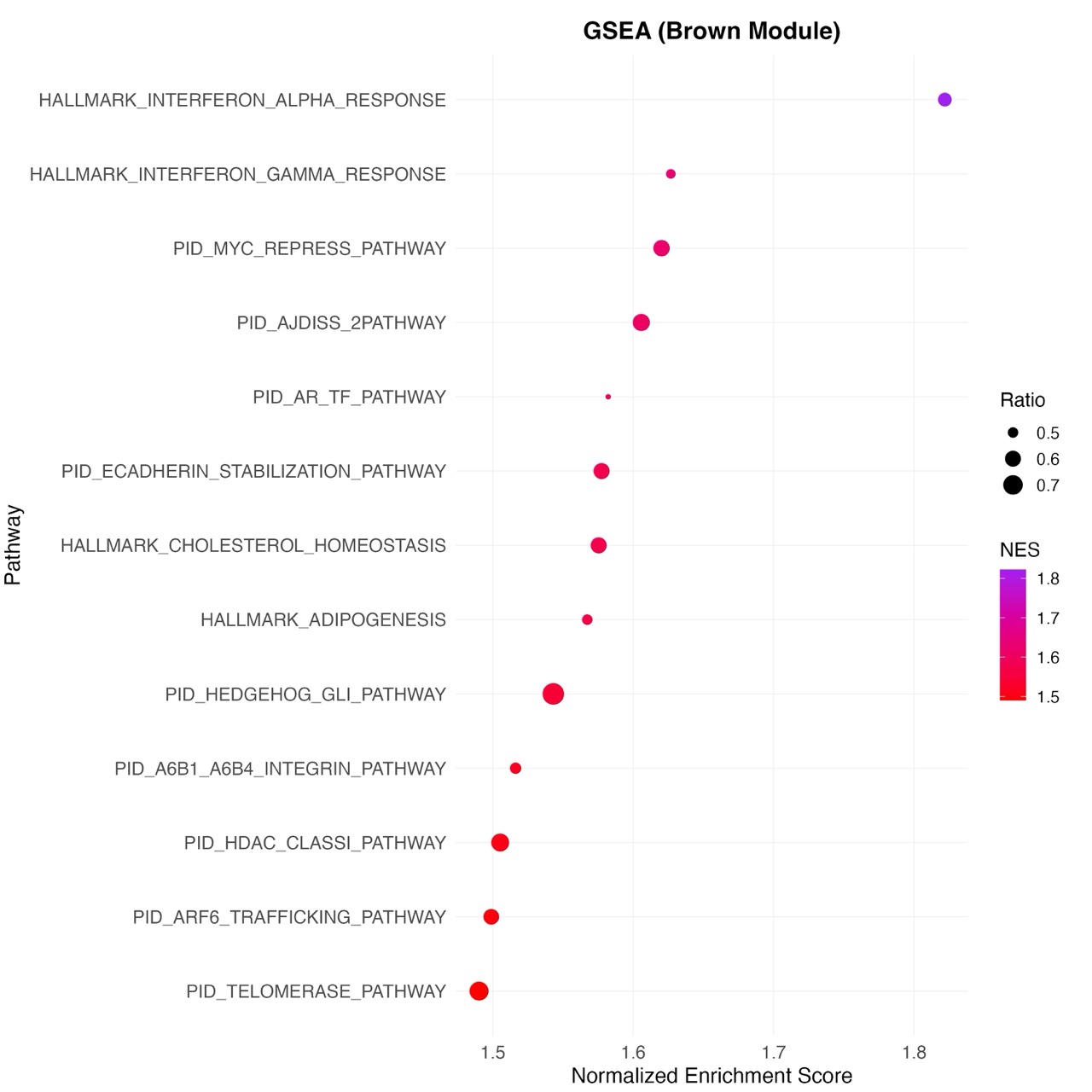
